# Vitamin D and socioeconomic deprivation mediate COVID-19 ethnic health disparities

**DOI:** 10.1101/2021.09.20.21263865

**Authors:** Leonardo Mariño-Ramírez, Maria Ahmad, Lavanya Rishishwar, Shashwat Deepali Nagar, Kara K. Lee, Emily T. Norris, I. King Jordan

## Abstract

Ethnic minorities in developed countries suffer a disproportionately high burden of COVID-19 morbidity and mortality, and COVID-19 ethnic disparities have been attributed to social determinants of health. Vitamin D has been proposed as a modifiable risk factor that could mitigate COVID-19 health disparities. We investigated the relationship between vitamin D and COVID-19 susceptibility and severity using the UK Biobank, a large progressive cohort study of the United Kingdom population. Structural equation modelling was used to evaluate the ability of vitamin D, socioeconomic deprivation, and other known risk factors to mediate COVID-19 ethnic health disparities. Asian ethnicity is associated with higher COVID-19 susceptibility, compared to the majority White population, and Asian and Black ethnicity are both associated with higher COVID-19 severity. Socioeconomic deprivation mediates all three ethnic disparities and shows the highest overall signal of mediation for any COVID-19 risk factor. Vitamin supplements, including vitamin D, mediate the Asian disparity in COVID-19 susceptibility, and serum 25-hydroxyvitamin D (calcifediol) levels mediate Asian and Black COVID-19 severity disparities. Several measures of overall health also mediate COVID-19 ethnic disparities, underscoring the importance of comorbidities. Our results support ethnic minorities’ use of vitamin D as both a prophylactic and a supplemental therapeutic for COVID-19.

## Introduction

The COVID-19 pandemic has had a disproportionate impact on ethnic minorities in developed countries^1-4^. In the United Kingdom (UK), Asian and Black minority groups have higher rates of COVID-19 diagnosis and mortality than the majority White population^5^. In the United States (US), American Indian, Black, and Latino populations have higher rates of COVID-19 cases, hospitalization, and death compared to White and Asian groups^6^. Ethnic disparities in COVID-19 have been attributed to a number of different factors, including a high burden of comorbidities, over-crowded living and working conditions, and reduced access to healthcare. Many of the risk factors associated with COVID-19 ethnic disparities are linked to structural barriers resulting from socioeconomic deprivation, segregation, and racial discrimination^7^.

The vitamin D hypothesis holds that low levels of circulating vitamin D among ethnic minority patients may also contribute to COVID-19 disparities^8^. Vitamin D plays an important role in a number of immune related processes, including defense against viral infections^9^. Biosynthesis of vitamin D depends on exposure to sunlight and ultraviolet irradiation of 7-dehydrocholesterol, which is converted to vitamin D_3_ (cholecalciferol) in the skin and then 25-hydroxyvitamin D (calcifediol) in the liver^10^. Melanin absorbs ultraviolet radiation, thereby reducing the amount of irradiation available for vitamin D synthesis in the skin. Accordingly, darker skinned individuals have reduced vitamin D biosynthesis, and lower levels of circulating 25-hydroxyvitamin D, than lighter skinned individuals^11^. This is particularly true for ethnic minorities living in high latitude countries like the UK^12^.

Studies on the role of vitamin D in immune defense against COVID-19 have yielded conflicting results. Retrospective cohort studies in the US found that vitamin D deficiency was associated with COVID-19 infection, with the strongest effect seen for Black patients^13,14^. A randomized control trial of hospitalized COVID-19 patients in Spain showed that treatment with a high dose of 25-hydroxyvitamin D significantly reduced admission to the intensive care unit^15^. A commentary in the British Medical Journal pointed to more than 40 such studies claiming that vitamin D mitigates COVID-19^16^. On the other hand, the UK National Institute for Health and Care Excellence surveyed the existing literature and concluded that there is insufficient evidence to support the use of vitamin D supplements for the treatment or prevention of COVID-19^17^. A pair of studies on the UK Biobank, a large progressive cohort study of the UK population, found that circulating levels of 25-hydroxyvitamin D are not significantly associated with the risk of COVID-19 disease or mortality^18,19^.

The objective of this study was to investigate the relationship between vitamin D and ethnic health disparities using the UK Biobank. The previous studies of the UK Biobank cohort relied on multivariable modelling to evaluate the association of vitamin D with COVID-19 outcomes when all other variables, including patient ethnicity, were held constant^18,19^. The multivariable modelling approach does not directly address which risk factors may explain ethnic differences in COVID-19 outcomes. For this study, we used structural equation modelling (SEM) to evaluate the ability of vitamin D, socioeconomic deprivation, and other known risk factors to mediate COVID-19 ethnic health disparities. The SEM approach allows for the formulation and testing of causal hypotheses with observational data as well as the characterization of potential intervening mechanisms. Our study cohort includes an order of magnitude greater sample size than the previous UK Biobank studies allowing for greater statistical resolution on the question of COVID-19 disparities.

## Methods

### Study cohort and participant attributes

The study cohort was obtained from the UK Biobank, a prospective cohort study intended to investigate the effects of genetic, environmental, and lifestyle factors on human health and disease^20^. The UK Biobank provides deep phenotypic, clinical, and genetic data for more than 500,000 participants enrolled between 2006 and 2010^21^.

We extracted the following information for UK Biobank participants: (1) age (Field 21022 Age at recruitment), (2) sex (Field 31: Sex), (3) ethnicity (Field 21000: ethnic background), (4) body mass index (Field 21001: Body mass index (BMI)), (5) smoking status (Field 20116: Smoking status), (6) diastolic blood pressure (Field 4079: Diastolic blood pressure, automated reading), (7) systolic blood pressure (Field 4080: Systolic blood pressure, automated reading), (8) Townsend deprivation index (Field 189: Townsend deprivation index at recruitment), (9) overall health rating (Field 2178: Overall health rating), (10) long-standing illness (Field 2188: Long-standing illness, disability, or infirmity), (11) supplements (Field 6155: Vitamin and mineral supplements), (12) vitamin D (Field 30890: Vitamin D), and (13) skin color (Field 1717: Skin colour).

UK biobank participants’ serum 25-hydroxyvitamin D (calcifediol) levels were measured in nmol/L units via chemiluminescent immunoassay analysis (CLIA) using the DiaSorin Liaison XL platform as previously reported (see Field 30890: Vitamin D description)^21,22^. Participants’ ages ranged from 40 to 69 at enrollment, and age at enrollment was split into decades for analysis. The categories for self-reported skin color (SRSC), which are Very fair, Fair, Light olive, Dark olive, Brown, and Black, were collapsed into four categories for analysis: Fair, Olive, Brown, and Black. Participant self-reported supplement use was considered for vitamins A, B, C, D, E, V9, and multivitamins. The effects of supplement use were modelled individually and jointly. Socioeconomic deprivation (SED) was measured by the Townsend deprivation index (TDI), a widely used measure of SED, which is known to be associated with poor health outcomes^23^. The TDI incorporates four factors into its score: (1) unemployment (as a percentage of those aged 16 and economically active), (2) non-car ownership (as a percentage of all households, (3) non-home ownership (as a percentage of all households), and (4) household overcrowding^24^. UK Biobank TDI values range from -6 to 11, where negative values in indicate less deprivation and higher values indicate more deprivation, and the raw values were split into deciles for analysis using the thresholds provided by the UK Biobank.

### Participant Ethnicity

UK Biobank participants self-identified as belonging to one of six ethnic groups at enrollment – Asian, Black, Chinese, Mixed, White, or Other – each of which is made up of a specific set of ethnic backgrounds. The UK Biobank ethnic group designations are based on the UK National Healthcare Service (NHS) categories, which were adopted from the UK Census. Since the UK Census makes no distinction between race and ethnicity, the UK Biobank ethnic groups used here may correspond to racial groups used in the US. The three largest ethnic groups in the UK Biobank – Asian, Black, and White – were used for analysis here. The Asian ethnic group consists of participants with Indian, Pakistani, Bangladeshi, or Any other Asian ethnic background. The Asian group does not include Chinese ethnicity, which has its own group. The Black ethnic group consists of participants with Caribbean, African, and Any other Black ethnic background. The White ethnic group consists of participants with British, Irish, and Any other White ethnic background.

### COVID-19 phenotypes and prevalence

Data on COVID-19 outcomes were taken from the UK Biobank covid19_result table (accessed December 20, 2020). COVID-19 susceptibility is defined as whether or not a participant tested positive for SARS-CoV-2, where cases are participants with positive test results and controls are participants with negative test results. SARS-CoV-2 PCR tests were performed on participant nose and throat swabs as previously described^25^. COVID-19 severity is defined as whether or not a participant with a SARS-CoV-2 positive test result was hospitalized, where cases are hospitalized participants with positive test results and controls are non-hospitalized participants with positive test results. Unadjusted prevalence values for COVID-19 susceptibility and severity were calculated as the number of cases divided by the number of cases and controls.

### Statistical analyses and visualization

All statistical analyses were performed using the R statistical language v4.0.4^26^. Bar graphs and heat maps were generated using the R ‘ggplot’ v3.3.3 library. The effects of ethnicity on COVID-19 susceptibility and severity (cases and controls) were modeled with age and sex as covariates using multivariable logistic regression with the ‘glm’ function in R: COVID-19 ∼ Ethnicity + Age + Sex. Forest plots showing the logistic regression coefficient point estimates (β-values), 95% confidence intervals, and P-values were generated using the ‘viz-forest’ function from the ‘meta-viz’ v0.3.1 package in R^27^.

Structural equation modelling (SEM) and mediation analyses were performed using the ‘sem’ function from the R statistical package ‘Lavaan’^28^. SEM was used to evaluate the ability of vitamin D, socioeconomic deprivation, and other known COVID-19 risk factors to mediate ethnic disparities in COVID-19 susceptibility and severity. Barron and Kenny’s stepwise method for mediation was used to evaluate the mediation effect of each risk factor independently, and then all risk factors that showed significant effects when considered independently were carried forward to a final multivariable structural equation model^29^. The ‘sem’ function was used to formulate each structural equation model and to calculate model coefficient values (β-values) and their P-values. For the final multivariable structural equation models, the mediation effects and their P-values were calculated using the ‘sem’ function. Mediation effects (indirect, direct, and total) are expressed as effect sizes (β-values), P-values, and percent mediation values. Structural equation path diagrams were generated using the web-based diagram tool ‘Lucidchart’. Additional details on the SEM approach used here are provided in the Supplementary Material.

## Results

### COVID-19 ethnic disparities

We characterized the prevalence of COVID-19 susceptibility and severity for the three largest ethnic groups in the UK Biobank: Asian, Black, and White. Demographic and ethnic characteristics for the study cohort are summarized in Table 1. Numbers are shown for the total cohort and for SARS-CoV-2 test positive cases and hospitalized cases, which were used to define the COVID-19 susceptibility and severity phenotypes. 35,339 study participants were tested for SARS-CoV-2, resulting in 6,049 positive cases, 1,630 of whom were hospitalized.

**Table 1.** Study cohort characteristics. Numbers and the percent share for each cohort category – total cohort, SARS-CoV-2 positive cases, and COVID-19 hospitalized cases – are shown for age, ethnicity, and sex.

| Characteristic | Total cohort<br>(n = 35,339) | Positive Cases<br>(n = 6,049) | Hospitalized Cases<br>(n = 1,630) |
| --- | --- | --- | --- |
| *Age – no. (cohort share %)* |  |  |  |
| < 45 | 3,392 (9.58%) | 1,076 (17.79%) | 155 (9.51%) |
| 45 – 54 | 8,994 (25.41%) | 2,123 (35.10%) | 390 (23.93%) |
| 55 – 64 | 14,607 (41.26%) | 1,849 (30.57%) | 598 (36.69%) |
| > 64 | 8,406 (23.75%) | 769 (12.71%) | 388 (23.80%) |
| *Mean age – year* | 57.37 | 54.11 | 57.99 |
| *Ethnic group – no. (cohort share %)* |  |  |  |
| Asian | 805 (2.27%) | 210 (3.47%) | 66 (4.05%) |
| Black | 670 (1.89%) | 137 (2.26%) | 54 (3.31%) |
| White | 33,924 (95.83%) | 5,702 (94.26%) | 1,510 (92.64%) |
| *Sex – no. (cohort share %)* |  |  |  |
| Female | 18,382 (51.93%) | 3,035 (50.17%) | 691 (42.39%) |
| Male | 17,017 (48.07%) | 3,014 (49.83%) | 939 (57.61%) |

The prevalence of COVID-19 susceptibility and severity were compared across ethnic groups, stratified by sex and age (Figure 1A). Males showed higher overall prevalence for COVID-19 susceptibility and severity compared to females. The prevalence of COVID-19 susceptibility decreases with age, whereas severity increases with age. The Asian ethnic group shows the highest prevalence for COVID-19 susceptibility, followed by the Black and White groups. The Black ethnic group shows the highest prevalence for COVID-19 severity, followed by the Asian and White groups.

**Figure 1.**
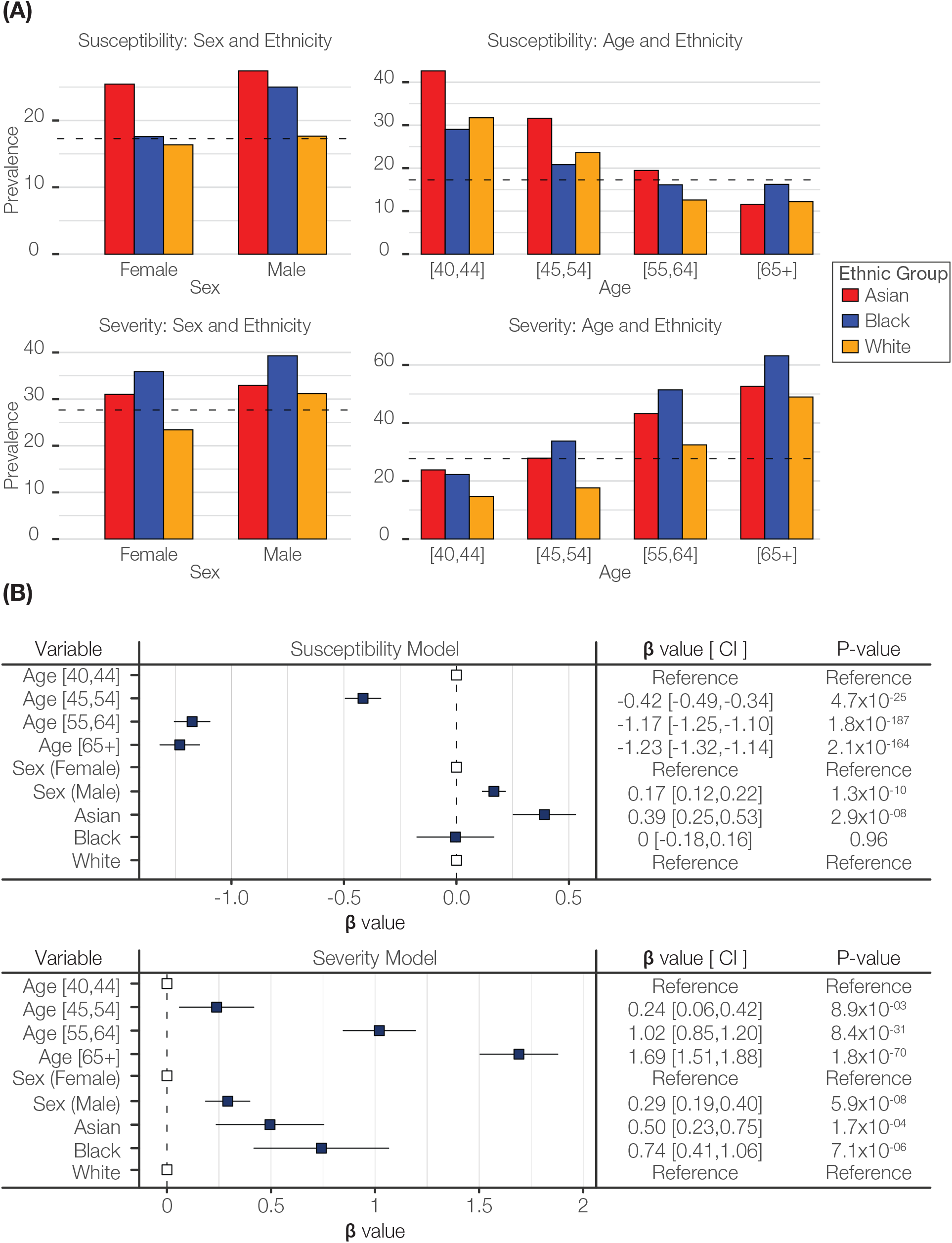
Ethnic disparities in COVID-19 susceptibility and severity. (A) Ethnic group prevalence values for COVID-19 susceptibility and severity: Asian (red), Black (blue), and White (orange). Prevalence values are stratified by sex and age brackets as shown. (B) Forest plots showing regression coefficient effect size estimates (β-values), 95% confidence intervals, and P-values for COVID-19 susceptibility and severity multivariable logistic regression models.

Multivariable logistic regression was used to model the joint effects of age, sex, and ethnicity on COVID-19 outcomes (Figure 1B). Age, sex, and ethnicity are all significantly associated with COVID-19 susceptibility and severity. Age shows the strongest effects on both susceptibility and severity, followed by sex and ethnicity. Asian ethnicity is significantly associated with higher susceptibility compared to the White reference group, whereas the Black and White groups do not show a significant difference in susceptibility. The Black and Asian groups both show significantly higher severity than the White reference group.

### Skin color, vitamin D, and socioeconomic deprivation

We explored the plausibility of the vitamin D hypothesis for COVID-19 disparities by looking at differences skin color and serum vitamin D levels among study participants from the Asian, Black, and White ethnic groups. Study participants from the Black ethnic group reported mainly black and brown skin color, and Asian participants reported primarily brown skin color, followed by olive and fair (Figure 2A). Participants from the White group reported mostly fair skin color, followed by olive.

**Figure 2.**
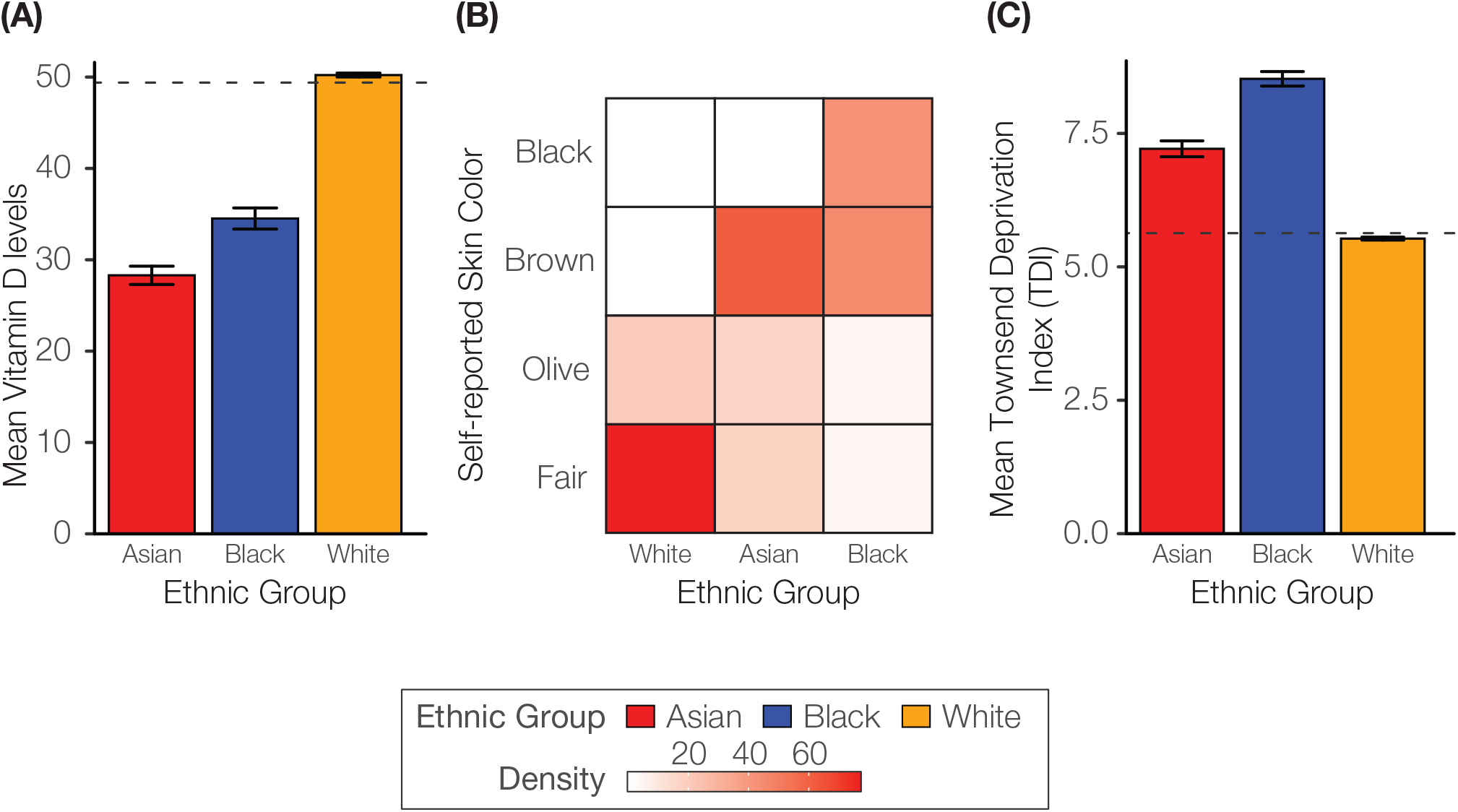
Ethnic differences in skin color, vitamin D levels, and socioeconomic deprivation. (A) Heatmap showing relative frequency (density) of participant self-reported skin color for Asian, Black, and White ethnic groups. (B) Mean ±95% confidence interval vitamin D levels for Asian (red), Black (blue), and White (orange) ethnic groups. (C) Mean ±95% confidence interval Townsend deprivation index (TDI) deciles for ethnic groups.

Participants from the White ethnic group showed the highest mean serum vitamin D levels, followed by the Black and Asian groups (Figure 2B). Differences in vitamin D levels between the White and other ethnic groups were much higher than the difference seen between the Black and Asian groups.

We also compared socioeconomic deprivation (SED) among ethnic groups using the composite Townsend deprivation index (TDI). The Black ethnic group shows the highest mean TDI decile followed by the Asian and White groups (Figure 2C).

### Testing the vitamin D hypothesis

We used structural equation modelling (SEM) to test the vitamin D hypothesis by evaluating the ability of vitamin D, and other known COVID-19 risk factors, to mediate the observed disparities in COVID-19 susceptibility and severity. In addition to vitamin D, we considered skin color, body mass index (BMI), systolic and diastolic blood pressure, long-standing illness (LSI), overall health ratio (OHR), smoking status, and supplement use as potential mediators of COVID-19 ethnic disparities. We were particularly interested in studying the role of socioeconomic deprivation, measured by the TDI, as a potential mediator of COVID-19 disparities. Age and sex were treated as exogenous variables with a direct effect on COVID-19 susceptibility and severity. We evaluated the mediation potential for each risk factor independently, and all risk factors that showed significant effects when considered alone were carried forward to a final multivariable structural equation model. This was done for each of the three significant COVID-19 ethnic disparities observed here: Susceptibility (Asian) and Severity (Asian and Black) (Figure 3). Each step in the SEM procedure is illustrated in Supplementary Figure 1 and the results for each step are shown in Supplementary Figures 2-5.

**Figure 3.**
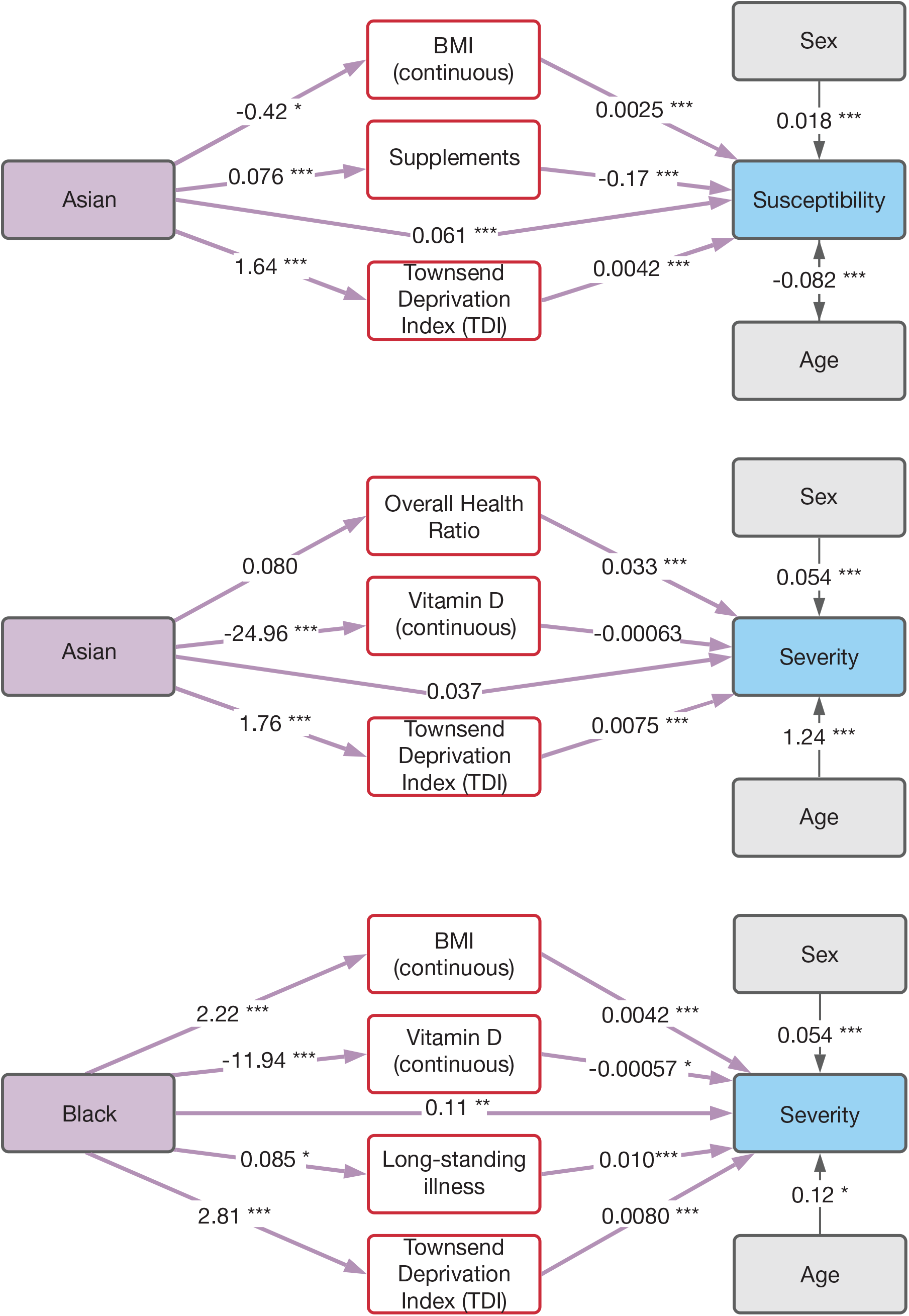
Mediation of COVID-19 ethnic disparities. Structural equation models are shown with ethnicity predictor variables (purple boxes), mediator variables (red boxes), exogenous predictor variables (gray boxes), and outcome variables (blue boxes). Arrows indicate the direction of predictor-to-outcome relationships. Direct effects of ethnicity on COVID-19 outcomes are shown along with the indirect (mediated) effects of ethnicity. Effect size estimates (β-values) are shown for each predictor-to-outcome relationship, and effect size statistical significance levels are indicated as *0.01<P≤0.05, **0.001<P≤0.01, ***P<0.001. Models are shown for Asian ethnicity and COVID-19 susceptibility (A), Asian ethnicity and COVID-19 severity (B), and Black ethnicity and COVID-19 severity (C). Detailed results for each of the three structural equation models are shown in Table 2.

The effect of Asian ethnicity on COVID-19 susceptibility is mediated by BMI, supplement use, and TDI, which shows the strongest mediating effect (Table 2). Supplement use includes vitamin D along with vitamins A, B, C, E, V9, and multivitamins. Together, the significant mediating factors contribute to a 14.6% indirect effect of Asian ethnicity on COVID-19 susceptibility. The direct effect of Asian ethnicity on COVID-19 susceptibility remains significant after mediation analysis.

**Table 2.** Mediation of COVID-19 ethnic disparities. Results of SEM mediation analysis are shown for Asian ethnicity and COVID-19 susceptibility (top), Asian ethnicity and COVID-19 severity (middle), and Black ethnicity and COVID-19 severity (bottom). Path diagrams for the three structural equation models are shown in Figure 3. Mediator variables, effect size estimates (β-values), significance levels (P-values), and effect percent mediation values are shown for each structural equation model.

| Variable | Effect size ($\beta$) | P-value | Effect (%) |
| --- | --- | --- | --- |
| **Effect of Asian ethnicity on COVID-19 susceptibility** |  |  |  |
| *Indirect (mediated) effects* |  |  |  |
| BMI | -1.0E-03 | 3.0E-02 | 1.4 |
| Supplements | -2.0E-03 | 1.0E-03 | 3.5 |
| TDI | 7.0E-03 | 1.2E-08 | 9.6 |
| *Total indirect effect* |  |  | 14.5 |
| *Direct effect* |  |  |  |
| Asian | 6.1E-02 | 3.6E-06 | 85.4 |
| *Total indirect + direct effect* |  |  |  |
|  | **6.5E-02** | **8.5E-07** | **100** |
| **Effect of Asian ethnicity on COVID-19 severity** |  |  |  |
| *Indirect (mediated) effects* |  |  |  |
| OHR | 2.1E-02 | 5.8E-05 | 29.3 |
| TDI | 1.3E-02 | 3.7E-04 | 14.1 |
| Vitamin D | 1.6E-02 | 1.5E-02 | 16.8 |
| *Total indirect effect* |  |  | 60.1 |
| *Direct effect* |  |  |  |
| Asian | 3.7E-02 | 0.2 | 39.9 |
| *Total indirect + direct effect* |  |  |  |
|  | **8.7E-02** | **3.7E-03** | **100** |
| **Effect of Black ethnicity on COVID-19 severity** |  |  |  |
| *Indirect (mediated) effects* |  |  |  |
| BMI | 9.0E-03 | 2.5E-03 | 5.8 |
| LSI | 9.0E-03 | 4.5E-02 | 5.4 |
| TDI | 2.3E-02 | 1.0E-04 | 14.3 |
| Vitamin D | 1.1E-02 | 3.1E-02 | 6.9 |
| *Total indirect effect* |  |  | 32.5 |
| *Direct effect* |  |  |  |
| Black | 0.1 | 4.1E-03 | 67.5 |
| *Total indirect + direct effect* |  |  |  |
|  | **0.2** | **1.7E-05** | **100** |

The effect of Asian ethnicity on COVID-19 severity is mediated by OHR, TDI, and vitamin D, and OHR shows the strongest mediating effect (Table 2). The three mediators contribute to a 60% indirect effect of Asian ethnicity on COVID-19 severity. The direct effect of Asian ethnicity on COVID-19 severity is no longer significant after mediation analysis, indicating complete mediation of this disparity.

The effect of Black ethnicity on COVID-19 severity is mediated by BMI, LSI, TDI, and vitamin D, and TDI shows the strongest mediating effect (Table 2). All mediators together contribute to a 32.5% indirect effect of Black ethnicity on COVID-19 severity. The direct effect of Black ethnicity on COVID-19 severity remains significant after mediation analysis.

## Discussion

### Mediation of COVID-19 ethnic disparities

Our analysis of the UK Biobank, a large prospective cohort study of the UK population, led to the identification of several risk factors that mediate ethnic disparities in COVID-19 susceptibility and severity. Our approach is distinguished from previous studies by the reliance on structural equation modeling (SEM), which allowed us to formulate and test causal hypotheses using the observational data available from the UK Biobank. Furthermore, our study cohort included an order of magnitude greater sample size than previous studies on vitamin D and COVID-19 in the UK Biobank as well as a more balanced case-control design that relied on negative SARS-CoV-2 tests as controls, as opposed to including non-tested individuals as controls.

The use of vitamin supplements was shown to mediate ethnic disparities in COVID-19 susceptibility but not severity. Body mass index (BMI), which is associated with numerous health outcomes, and two other metrics of overall health – long-standing illness (LSI) and overall health ratio (OHR) and – were shown to mediate COVID-19 ethnic disparities, consistent with an important role for comorbidities in elevating the risk of COVID-19 in minority patients.

Several known COVID-19 risk factors did not show any significant mitigating effects on COVID-19 disparities, including smoking, systolic and diastolic blood pressure. In contrast to the predictions of the vitamin D hypothesis for COVID-19 ethnic disparities, skin color did not show any significant mediating effect. This may be because skin color is self-reported in the UK Biobank, which could lead to reporting biases, and participants are asked to choose from several broad categories of color as opposed to using measurement of more continuous values.

### Efficacy of socioeconomic deprivation compared to vitamin D intervention

We report evidence that vitamin supplements, including vitamin D, mediate Asian disparity in COVID-19 susceptibility, and serum vitamin D levels mediate Asian and Black ethnic disparities in COVID-19 severity. However, socioeconomic deprivation, as measured by the Townsend deprivation index (TDI), was the only single risk factor that showed significant mediation in all three structural models, for both COVID-19 susceptibility and severity. In addition, the mediating effects of TDI on COVID-19 ethnic disparities were greater than seen for vitamin D. TDI had the highest mediating effect in two out of three structural models (Asian susceptibility and Black severity) and the second highest mediating effect in the third model (Asian severity).

Considered together, these results show that socioeconomic deprivation has the greatest impact on COVID-19 outcomes, and policies aimed at eliminating socioeconomic deprivation would do the most to mitigate COVID-19 ethnic disparities. However, policies of this kind are difficult to implement and could take generations to realize their effects. Vitamin D, on the other hand, is a modifiable risk factor that is inexpensive, widely available, and easy to administer. Furthermore, vitamin D has a number of health benefits beyond the effects on COVID-19 measured here. Given the finding that Asian and Black individuals in the UK have lower levels of circulating vitamin D, we recommend that ethnic minorities take 25-hydroxyvitamin D (calcifediol) supplements as a routine method of preventative healthcare.

### Limitations of the study and future directions

Our evaluation of the role of vitamin D in COVID-19 ethnic disparities is limited by the reliance on observational data from the UK Biobank. As discussed previously, the SEM approach allowed us to formulate and test hypotheses using observational data regarding the causes of COVID-19 disparities and to characterize potential intervening mechanisms that could be used to mitigate the disparities. However, our results could be effected by unobserved confounders, and randomized control trials are the gold standard for definitely proving the efficacy of any medical intervention. Nevertheless, our results can be taken to support the establishment of clinical trials on the efficacy of vitamin D as a prophylactic and treatment for COVID-19, with an emphasis on the recruitment of participants from ethnic minority groups.

## Data Availability

Data are made publicly available by the UK Biobank.

