## Supplementary Material for "Vitamin D and socioeconomic deprivation mediate COVID-19 ethnic health disparities"

### *Structural equation modelling and mediation analysis*

We used Barron and Kenny's steps for mediation to evaluate the ability of vitamin D, socioeconomic deprivation, and other known COVID-19 risk factors to mediate ethnic disparities in COVID-19 susceptibility and severity. Path diagrams corresponding to the five steps used for Barron and Kenny's mediation analysis are shown in Supplementary Figure 1, and the steps are explained below. For each step in the procedure, effect sizes (β-values) and their significance levels (P-values) were calculated.

Step #1: X predicts Y. Calculate the effects of the predictors (X) on the outcomes (Y). The predictors are ethnicity and the outcomes are COVID-19 susceptibility or severity. Age and sex are included as exogenous covariates that can affect the outcomes.

Step #2: X predicts M. Calculate the effects of the predictors (X) on the mediators (M). The predictors are ethnicity and the mediators are the COVID-19 risk factors. Age and sex are included as exogenous covariates that can affect the mediators.

Step #3: M predicts Y. Calculate the effects of the mediators (M) on the outcomes (Y). The mediators are the COVID-19 risk factors and the outcomes are COVID-19 susceptibility or severity. Age and sex are included as exogenous covariates that can affect the outcomes.

Predictor and mediator variables that showed statistically significant effects, when considered independently, in steps #1-#3 were carried forward to step #4. In step #4, individual predictors (ethnicity) and individual mediators are considered together to measure direct effects of the predictors on the outcomes and indirect effects of the predictors on the outcomes mediated by the mediators.

Step #4: X and M predict Y. Calculate the direct effects of the predictors (X) on the outcomes (Y), and calculate the indirect effects of the predictors (X) on the outcomes (Y) that are mediated by the mediators (M). The predictors are ethnicity, the outcomes are COVID-19 susceptibility or severity, and the mediators are the COVID-19 risk factors. The indirect effects of the mediators (M) are calculated as the product of the effect sizes (β-values) for X -> M and M-> Y. Age and sex are included as exogenous covariates that can affect the outcomes.

If both the direct and indirect of paths showed statistically significant effects on the outcome in step #4, the predictor (X, ethnicity) and the mediator (M, COVID-19 risk factor) were carried forward to the final multivariable structural equation model for each disparity where the effects of multiple mediators are considered together.

Step #5: X and multiple M predict Y. Calculate the direct effects of the predictors (X) on the outcomes (Y), and calculate the indirect effects of the predictors (X) on the outcomes (Y) that are mediated by multiple mediators (M). The predictors are ethnicity, the outcomes are COVID-19 susceptibility or severity, and the mediators are the COVID-19 risk factors. The indirect effects of the mediators (M) are calculated as the product of the effect sizes (β-values) for X -> M and M-> Y. Age and sex are included as exogenous covariates that can affect the outcomes. Final multivariable models and results for step #5 are shown in Figure 3 and Table 2 in the main body of the manuscript. For the final multivariable structural equation models, the mediation effects (indirect, direct, and total) are expressed as effect sizes (β-values), P-values, and percent mediation values (see Table 2 in the main body of the manuscript).

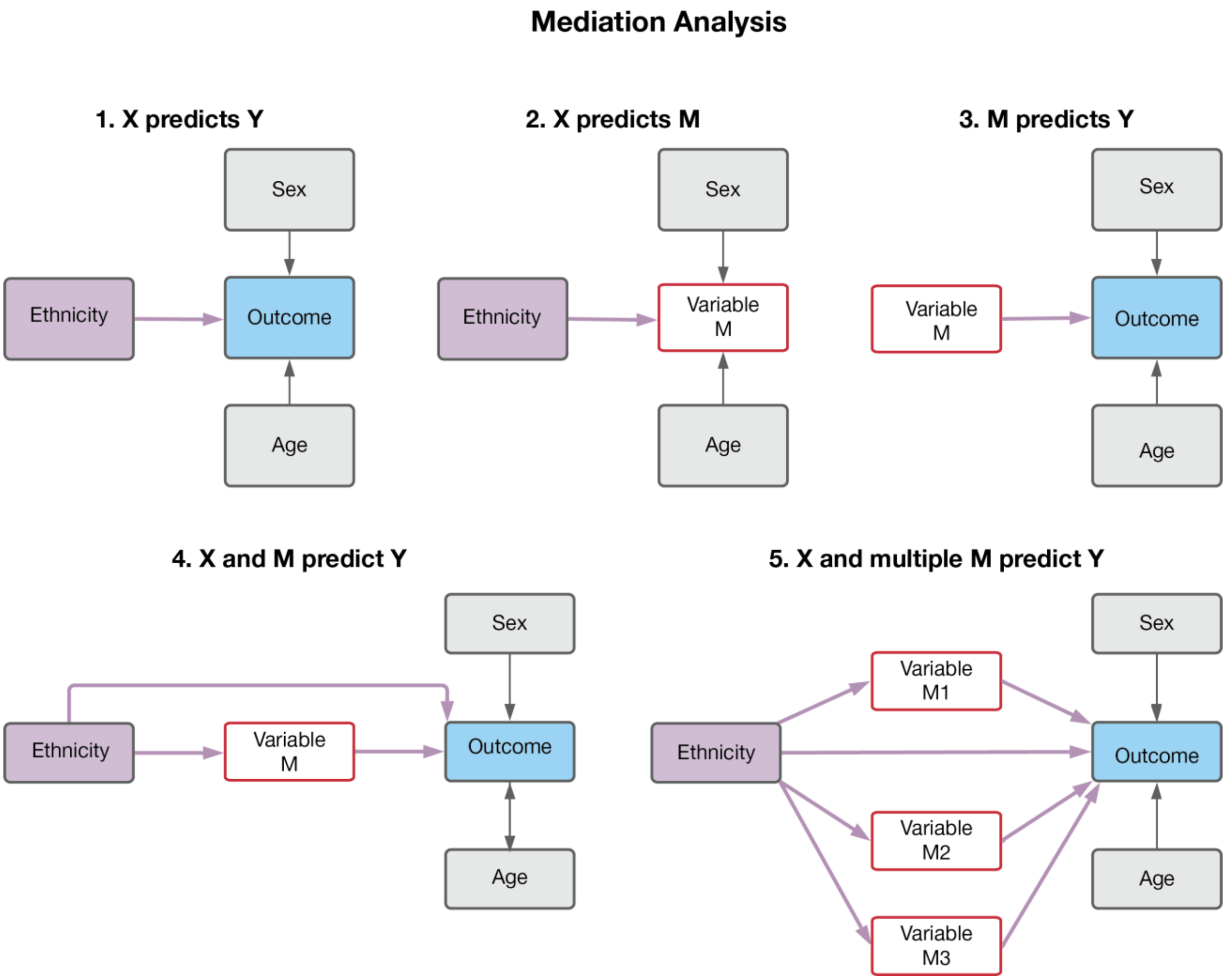


Supplementary Figure 1. **Structural equation models used for COVID-19 ethnic disparity mediation analysis.** Models are shown as path diagrams with model variables in boxes and arrows indicating the relationship between independent (predictor) variables and dependent (outcome) variables. The five steps in the Barron and Kenny's stepwise method for mediation are described in detail above. The results for each step in the analysis are shown in Supplementary Figures 2-6.

| COVID-19 Susceptibility | | COVID-19 Severity | |
|---|---|---|---|
| Asian | Black | Asian | Black |
| 0.065 | 0.089 | 0.089 | 0.162 |

Supplementary Figure 2. **Results for step #1 of Barron and Kenny's stepwise method for mediation.** Ethnicity (X) predicts COVID-19 susceptibility or severity (Y) (Supplementary Figure 1.1). Effect size estimates (β-values) are shown for ethnicity predictors. Statistically significant effect sizes (P<0.05) are indicated with green shading.

| Outcome | Ethnicity | Total effect | Exogenous Variables | | Mediators | | | | | | | | | |
|---|---|---|---|---|---|---|---|---|---|---|---|---|---|---|
| | | | Age | Sex | BMI | Diastolic BP | LSI | OHR | Smoking | SRSC | Supp. | Systolic BP | TDI | Vitamin D |
| COVID-19 Susceptibility | Asian | ~ | ~ | ~ | -0.37 | -0.23 | -0.02 | 0.08 | 0.00 | 0.60 | 0.09 | -1.30 | 1.57 | -21.56 |
| | Black | | | | | | | | | | | | | |
| COVID-19 Severity | Asian | ~ | ~ | ~ | -0.20 | -1.57 | 0.05 | 0.09 | -0.03 | 0.55 | 0.11 | -1.60 | 1.76 | -24.80 |
| | Black | ~ | ~ | ~ | 2.35 | 3.80 | 0.12 | 0.05 | -0.01 | 0.73 | 0.16 | 6.80 | 2.81 | -19.26 |

Supplementary Figure 3. **Results for step #2 of Barron and Kenny's stepwise method for mediation.** Ethnicity (X) predicts COVID-19 mediators (M) (Supplementary Figure 1.2). Effect size estimates (β-values) are shown for ethnicity predictors. Statistically significant effect sizes (P<0.05) are indicated with green shading. Mediator abbreviations: BMI – body mass index, BP – blood pressure, SRSC – self-reported skin color, Supp. – supplement use (Vitamin A, B, C, D, E, V9, Multivitamin), TDI – Townsend deprivation index.

| Outcome | Ethnicity | Total effect | Exogenous Variables | | Mediators | | | | | | | | | |
|---|---|---|---|---|---|---|---|---|---|---|---|---|---|---|
| | | | Age | Sex | BMI | Diastolic BP | LSI | OHR | Smoking | SRSC | Supp. | Systolic BP | TDI | Vitamin D |
| COVID-19 Susceptibility | Asian | ~ | ~ | ~ | 0.003 | 0 | -0.014 | 0.013 | 0.032 | 0.062 | -0.169 | 0 | 0.005 | 0 |
| | Black | | | | | | | | | | | | | |
| COVID-19 Severity | Asian | ~ | ~ | ~ | 0.01 | 1.4E-04 | 0.12 | 0.37 | -0.1 | 0.63 | 0.2 | 3.6E-04 | 0.01 | -0.001 |
| | Black | ~ | ~ | ~ | 0.01 | 5.7E-04 | 0.12 | 0.39 | -0.09 | 0.24 | 0.19 | 5.2E-04 | 0.01 | -0.001 |

Supplementary Figure 4. **Results for step #3 of Barron and Kenny's stepwise method for mediation.** Mediators (M) predict COVID-19 susceptibility or severity (Y) (Supplementary Figure 1.3). Effect size estimates (β-values) are shown for mediator predictors. Statistically significant effect sizes (P<0.05) are indicated with green shading.

| Outcome | Ethnicity | Total effect | Exogenous Variables | | Mediators | | | | | | | | | |
|---|---|---|---|---|---|---|---|---|---|---|---|---|---|---|
| | | | Age | Sex | BMI | Diastolic BP | LSI | OHR | Smoking | SRSC | Supp. | Systolic BP | TDI | Vitamin D |
| COVID-19 Susceptibility | Asian | 0.065 | -0.08 | 0.02 | -1.20E-03 | 1.50E-05 | 5.90E-04 | 3.20E-04 | -3.90E-03 | -4.60E-03 | -2.30E-03 | -1.70E-04 | 7.40E-03 | -5.60E-03 |
| | Black | | | | | | | | | | | | | |
| COVID-19 Severity | Asian | 0.088 | 0.13 | 0.06 | -2.20E-03 | -3.00E-04 | 1.10E-03 | 2.30E-02 | -9.40E-03 | 9.60E-05 | 1.40E-03 | -1.30E-03 | 1.90E-02 | 2.60E-02 |
| | Black | 0.160 | 0.13 | 0.06 | 0.0151 | 0.0016 | 0.01 | 0.0121 | -0.0066 | -0.0515 | 0.0062 | 0.0022 | 0.0317 | 0.0199 |

Supplementary Figure 5. **Results for step #4 of Barron and Kenny's stepwise method for mediation.** Ethnicity (X) predicts mediators (M), which predict COVID-19 susceptibility or severity (Y) (Supplementary Figure 1.4). Total effect size estimates (β-values) for ethnicity and indirect effect size estimates for mediators are shown. Statistically significant effect sizes (P<0.05) are indicated with green shading.
